# NOVEL SOLUTION BASED ON DETECTION OF MIRS-410-3P AND 141-5P FOR DIAGNOSTIC OF PROSTATE CANCER EVOLUTION

**DOI:** 10.1101/2024.03.11.24303774

**Authors:** M.F. Carbache, Y. Ouahid, E. Sainz, J.J. Montoya, A. Molera, J. Martínez-Olmos, D. Cortés, W. Golomar, J. Carballido, P. Castán

## Abstract

Prostate cancer (PCa) remains the most frequently diagnosed malignancy in men and the second leading cause of cancer-related mortality worldwide. Population-based prevention and screening programmes have improved early detection rates; however, current diagnostic pathways—largely driven by prostate-specific antigen (PSA) testing and imaging—still suffer from limited sensitivity and specificity. This limitation contributes to substantial rates of unnecessary biopsies, overdiagnosis and overtreatment, highlighting the need for more accurate molecular stratification tools.

In recent years, circulating microRNAs (miRNAs) have emerged as promising biomarkers for cancer diagnosis, risk reclassification, prediction of tumour progression and treatment response. Among them, miR-410-3p and miR-141-5p have been consistently implicated in prostate cancer biology. Previous studies in tumour tissues and prostate cancer cell lines have demonstrated that elevated miR-410-3p expression correlates with discordant clinical scenarios in which PSA levels fail to match biopsy, surgical or digital rectal examination findings, as well as with poor patient prognosis. In parallel, miR-141-5p exhibits complementary behaviour, supporting the rationale for a dual-biomarker approach based on relative expression patterns rather than single-marker quantification.

Mechanistically, miR-410-3p has been shown to exert oncogenic activity through downregulation of PTEN, leading to activation of the AKT/mTOR signalling pathway. Notably, divergent expression dynamics of miR-410-3p have been reported between tumour tissues, cancer cell lines and peripheral blood, reinforcing the value of combined assessment with miR-141-5p. Large cohort studies (n > 500) have confirmed upregulation of miR-141-5p in prostate cancer patients at both epithelial and stromal levels, while concomitant reduction of circulating miR-410-3p has been associated with increased risk of biochemical recurrence.

In this study, we present the design, molecular configuration and preclinical evaluation of a novel RTqPCR-based diagnostic system that leverages residual blood volumes routinely discarded after PSA testing. The system enables semi-quantitative, parallel detection of miR-410-3p and miR-141-5p in plasma, providing a non-invasive molecular readout of prostate cancer progression and recurrence risk. The results support the feasibility of this approach as a complementary diagnostic tool with the potential to reduce reliance on invasive procedures such as biopsy, surgery and digital rectal examination, while improving molecular precision in prostate cancer management.

## Introduction

Prostate cancer (PCa) is currently one of the most extensively characterised solid tumours from a molecular standpoint. Advances in transcriptomics and post-transcriptional regulation have progressively delineated a complex regulatory landscape in which non-coding RNAs—particularly microRNAs (miRNAs)—play a pivotal role in tumor initiation, progression, and metastatic dissemination. Over the last decade, a growing body of evidence has established specific miRNA signatures as informative biomarkers for diagnosis, prognosis, and disease monitoring in PCa. Among these, miR-410-3p and miR-141-5p have emerged as two of the most consistently reported and biologically relevant candidates, exhibiting complementary and stage-dependent behaviour that makes them particularly attractive for combined diagnostic strategies (8,9).

MiRNAs are a class of small, non-coding RNA molecules that exert post-transcriptional control of gene expression through sequence-specific binding to the 3′ untranslated region (3′-UTR) of target mRNAs. This interaction recruits the Argonaute (Ago) protein within the RNA-induced silencing complex (RISC), leading to translational repression or mRNA degradation (10). Through this mechanism, miRNAs function as high-level regulatory nodes capable of modulating entire signalling pathways rather than isolated genes. In oncological contexts, dysregulation of miRNA expression can therefore drive phenotypic shifts associated with uncontrolled proliferation, resistance to apoptosis, invasion, and metastatic competence.

A particularly compelling feature of miRNAs is their detectability in peripheral biofluids, including plasma and serum, where they display remarkable stability despite the presence of endogenous RNases. This property, combined with their functional involvement in oncogenic processes, positions circulating miRNAs as powerful tools for non-invasive cancer diagnostics, risk stratification, and longitudinal disease monitoring. Indeed, variations in circulating miRNA levels have been shown to correlate with tumour burden, molecular subtype, treatment response, and clinical outcome across multiple cancer types, including PCa (4).

Within this framework, miR-410-3p has been extensively characterised as a key oncogenic miRNA in prostate cancer biology. The seminal work by Zhang et al. (1) demonstrated that elevated expression of miR-410-3p is associated with downregulation of the tumour suppressor phosphatase and tensin homolog (PTEN), resulting in constitutive activation of the AKT/mTOR signalling pathway—a central driver of prostate carcinogenesis and therapeutic resistance (11). Complementary findings by Liu et al. (2) further established that miR-410-3p upregulation enhances tumour progression and metastatic potential in PCa through PTEN inhibition, reinforcing its role as a functional oncogene within the PI3K/AKT/mTOR axis. Notably, similar oncogenic behaviour of miR-410-3p has been observed in clear cell renal cell carcinoma (CCRCC), underscoring the broader relevance of this regulatory circuit across urological malignancies (12).

Beyond correlative expression data, functional studies provide compelling mechanistic validation. In vitro inhibition of miR-410-3p in widely used prostate cancer cell lines such as PC3 and DU145 results in significant suppression of cell proliferation, cell-cycle progression, migration, invasion, and tumour growth in vivo, while simultaneously promoting apoptotic pathways. These findings position miR-410-3p not merely as a passive biomarker, but as an active driver of malignant behaviour. Clinically, elevated miR-410-3p expression has been identified as an independent prognostic factor associated with poor survival outcomes in CCRCC and PCa, further reinforcing its translational significance (13–15). Mechanistic studies in additional tumour models, including lung cancer, have consistently confirmed that the oncogenic effects of miR-410-3p are mediated through PTEN suppression and downstream AKT/mTOR activation, highlighting the robustness and cross-context validity of this pathway (15).

In parallel, miR-141-5p—belonging to the miR-200 family—has been repeatedly linked to prostate cancer progression, particularly in advanced and metastatic disease stages. Large cohort studies exceeding 500 patients have demonstrated that elevated miR-141 expression is associated with increased risk of biochemical recurrence, biochemical failure, and clinical progression following primary treatment (10). Circulating miR-141-5p levels are significantly higher in patients with aggressive PCa phenotypes, including those with high Gleason scores and lymph-node involvement, suggesting its value as a marker of disease severity rather than early initiation alone.

Importantly, the dynamic and sometimes opposing behaviour of miR-410-3p and miR-141-5p across different stages of prostate cancer progression highlights a key limitation of single-biomarker strategies. While miR-141-5p tends to increase with tumour advancement, several studies have reported a relative decrease in circulating miR-410-3p levels in locally advanced disease compared with earlier stages (10,13,14). This reciprocal pattern provides a compelling biological rationale for a dual-marker approach, in which the combined assessment of miR-410-3p and miR-141-5p functions as a molecular cross-checkpoint, enhancing diagnostic resolution and reducing ambiguity associated with stage-specific expression shifts.

Supporting this strategy, both miRNAs have been reliably detected in liquid biopsy matrices— including urine, serum, plasma, and whole blood—demonstrating their ability to enter systemic circulation and be quantitatively measured using standard molecular techniques (16). The convergence of biological relevance, detectability, and complementary behaviour positions the miR-410-3p / miR-141-5p pair as one of the most promising biomarker sets for prostate cancer diagnosis and progression assessment currently described.

In this context, the present work introduces a novel RT-qPCR-based assay designed for the semi-quantitative, simultaneous detection of miR-410-3p and miR-141-5p. The system has been developed to maximise analytical robustness while remaining compatible with routine clinical workflows. Preclinical performance data are presented alongside analyses conducted on primary plasma samples obtained from patients attending the Urology Service of Hospital Universitario Puerta de Hierro, using discarded plasma volumes from routine PSA testing. This approach ensures ethical proportionality while directly anchoring the assay in real-world clinical material.

The novel platform, MiRNAX Biosens ProstREACT©, demonstrates concordance between in vitro validation data and results obtained from clinical specimens, supporting its potential translational value. Collectively, these findings suggest that this dual-miRNA assay may represent a promising non-invasive alternative or complement to first-line invasive diagnostic procedures such as prostate biopsy, digital rectal examination, or early surgical intervention, particularly in scenarios where PSA-based screening yields equivocal results.

## MATERIALS AND METHODS

sults. Between 2022 and 2023, 43 patients with a high probability of PCa or confirmed tumor and a positive PSA enzyme linked immunosorbent assay test (ELISA) (>4 ng/mL) were recruited in the Urology Service of Hospital Puerta de Hierro (Madrid) and re-tested independently at the MBR&DU (see filiation) to confirm the levels of MiR-410-3p and miR-141-5p on the novel system (MiRNAX Biosens ProstREACT©). The recruitment was done under the framework named “clinical proof of concept (PCC) of the cross-sectional analysis system of miRs for the exclusion of prostate inflammatory processes and diagnosis of cancer associated with PSA variations” supported by the team formed by MiRNAX Biosens S. L. and the Urology Department of the Hospital Puerta Hierro represented by its Head of Service (Dr. Joaquín Carballido), which was approved by the Ethics Committee for Clinical Research of the Research with Medicines of the Hospital Universitario Puerta de Hierro Majadahonda, Spain (supplemental documentation available from the authors).

Results were contrasted blindly with the PCa diagnostic formerly defined in accordance with the existing clinical guidelines. For this, patients underwent a biopsy test for prostate to confirm the cancer and the Gleason score was obtained in parallel to evaluate the microscopic features of the phenomenon found. Serum PSA was used as putative biomarker of PCa-associated risk and treatment prognosis. Afterwards, risk stratification was evaluated according to the following groups defined by the Urology Service in compliance with the clinical guidelines. During the period, 43 patients were diagnosed to have PCa, which complied with the ethical and technical requirements for the study set this research (documentation available through the Urology Service and Ethics Committee of Hospital Puerta de Hierro). The controls included healthy volunteers and all attendants are listed in the table below:

**Table 1.**
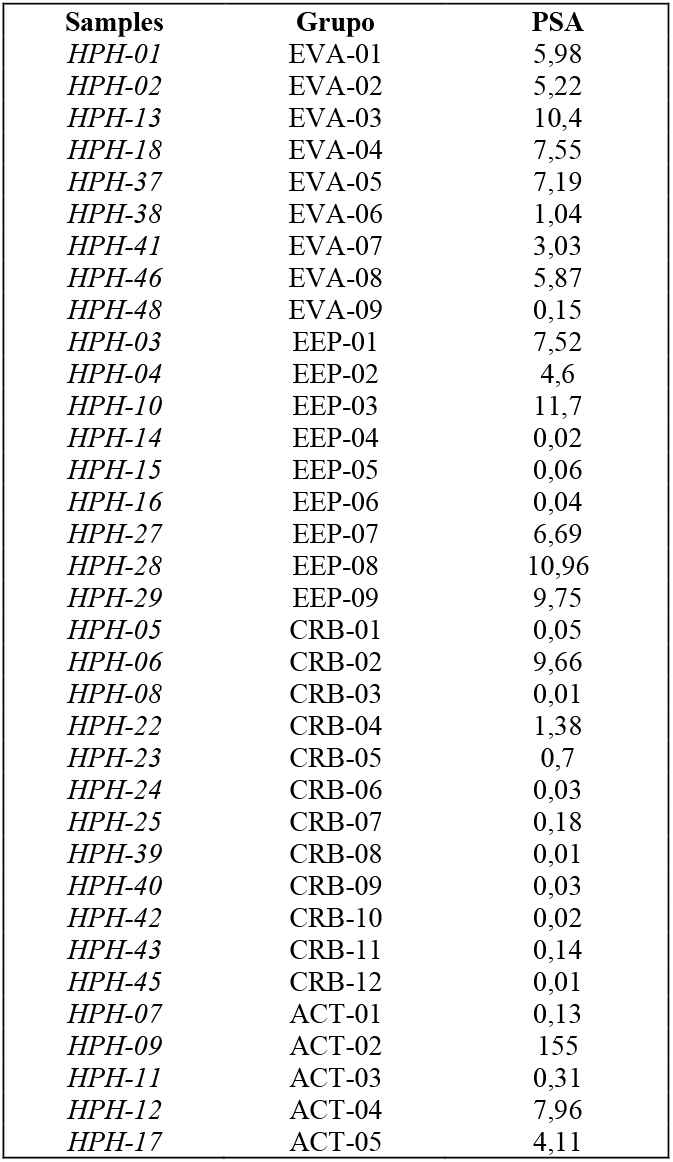
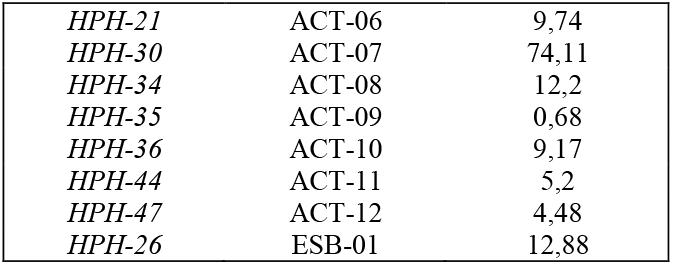
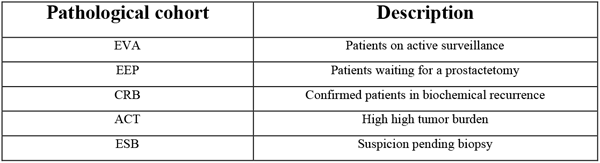
List of attendants diagnosed as per the existing clinical guidelines and tested for MiR-410-3p and miR-141-5p on the novel system (MiRNAX Biosens ProstREACT©).

### Serum sampling and total RNA isolation

Peripheral blood samples from donors were obtained at the Urology Service of Hospital Puerta de Hierro and allowed to stand for 1 hour before plasmas were obtained by separation from sera removing clots by centrifugation at room temperature for 10 minutes. The resulting supernatant (serum) was carefully removed to obtain the plasma from the whole blood samples collected in tubes with an anticoagulant (Vacutainer® 143425 EAN: 30382903688150 BD, New York, USA) as soon as the two phases were clearly observed. Resulting upper plasma aliquots (volumes ranging from 500 uL to 1 ml) were transferred to sterile 1.5 mL Eppendorf tubes labeled as “Primary Plasma” and stored at -80ºC. The remaining blood devoid of clot was thoroughly homogenized by gentle shaking and added to Falcon® tubes (Thermo-Fisher Scientific Ref:10788561-BD, New York, USA) containing 5 mL of Ficoll. Addition was carried very slowly, avoiding a turbulent blood-Ficoll interaction while allowing the carbohydrate and metrizamide polymer to cause precipitation of erythrocytes and granulocytes, maintaining mononuclear cells afloat. Once the process was completed and cell separation due to the difference of densities was visually evident, a second centrifugation was performed at 2500 rpm for 30 min at room temperature yielding 4 phases: Surface Secondary Plasma (SSP), Peripheral Blood Mononuclear Cells (PBMC), Ficoll and Erythrocytes. SSP was then recovered (volumes ranging from 500 uL to 1 ml) and transferred to a 1.5 mL Eppendorf tube labeled as “Secondary Plasma” before storing at -80ºC. Samples thus obtained were kept frozen until cohorts were completed before progressing to manipulate for the detection of miR-410-3p -5p and miR-141-5p. For this, total RNA was isolated using the miRNEasy® kit (Ref: 217184 Qiagen, USA) according to the manufacturer’s instructions for use. Briefly, 200 μL plasma aliquots were used to perform total RNA isolation. Total RNA was eluted in 14 μL of RNAse-free water and subsequent RNA quantification was performed without freezing using a NanoDrop® (Ref:ND-2000) recording the averaged absorbance at 230, 260, 280 and 330 nm from three reads to calculate the concentration and purity of the purified products.

Once the concentration values were recorded, all RNA samples were analyzed for RNA integrity using MiRNAX-Biosens Kit for determination of hRNAse-P expression® (Ref. M001) which uses a stable endogenous reference small coding RNA used to accurately assess the yield of intact RNA fit for further amplification within the total RNA pool isolated. This system provides an estimation based on the Ct criteria with valid samples registering amplification in the range of 21-26 Ct. The Ct values were then used to make sure that proportionate amounts of primary plasma borne intact RNA were used in each reaction for reverse transcription and qPCR.

**Figure 1.**
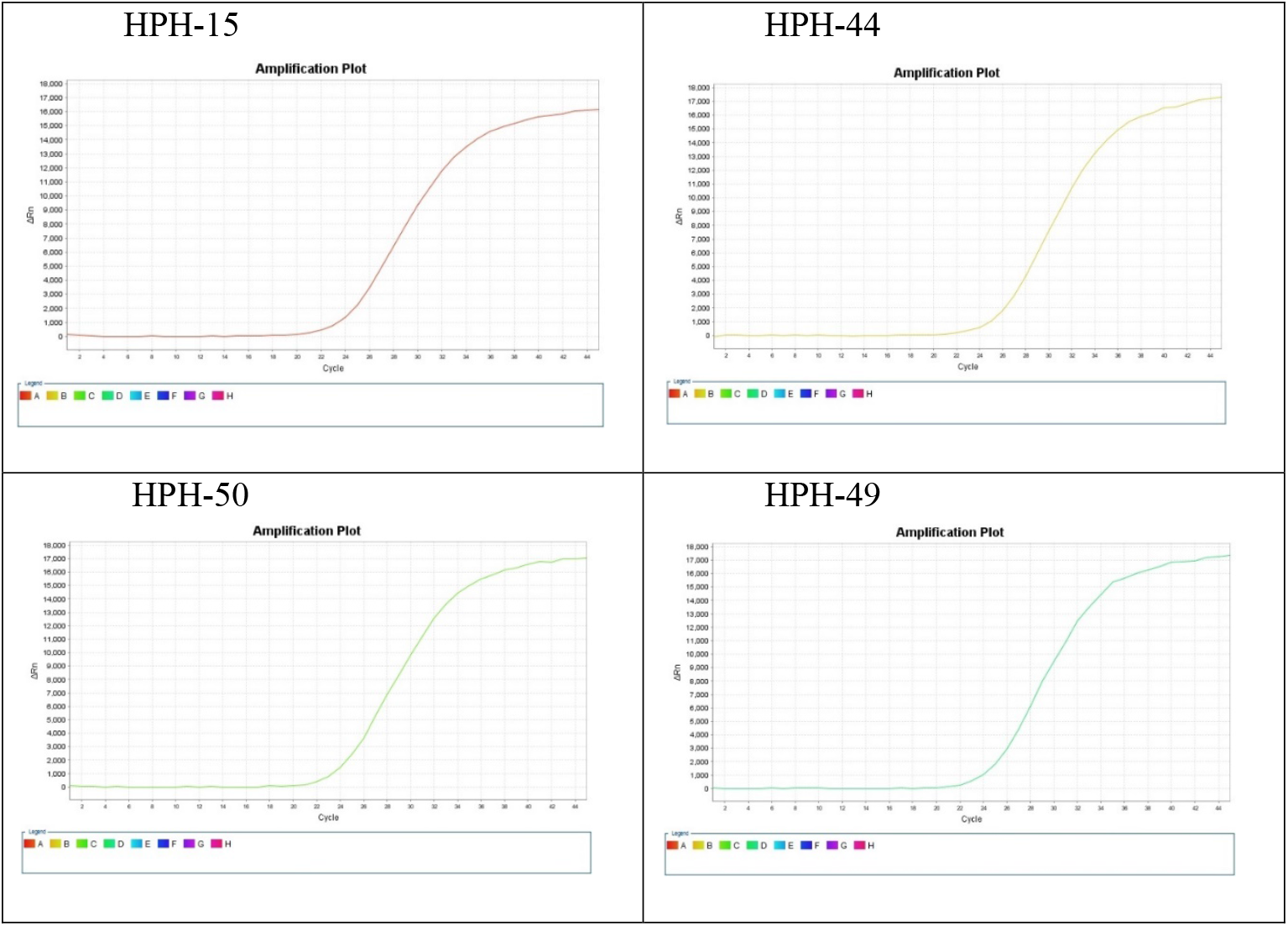
Example of primary and secondary plasmas analyzed for RNA integrity using MiRNAX-Biosens Kit for determination of hRNAse-P expression® (Ref. M001) for normalization as per the description above.

### Quantification of miR-410-3p and miR 141-5p expression and statistical analysis

MiR-410-3p and miR-141-5p were detected using the novel system (MiRNAX Biosens ProstREACT©), a patented RtqPCR assay (Patent Ref: EP4055189A1(17) ) for the semi quantitative detection of both biomarkers based on the staggered priming of the forward and reverse oligonucleotides building an extended amplicon fit for perfect-match hybridization of a FAM-MGB fluorescent probes covering the entire MiR-410-3p and miR-141-5p mature sequences. The specific primers for amplification of MiR-410-3p and miR-141-5p in this assay were subjected to in silico analysis for specificity and cross-reactivity before confirming the analytical sensitivity of the test (see Results).

**Figure 2.**
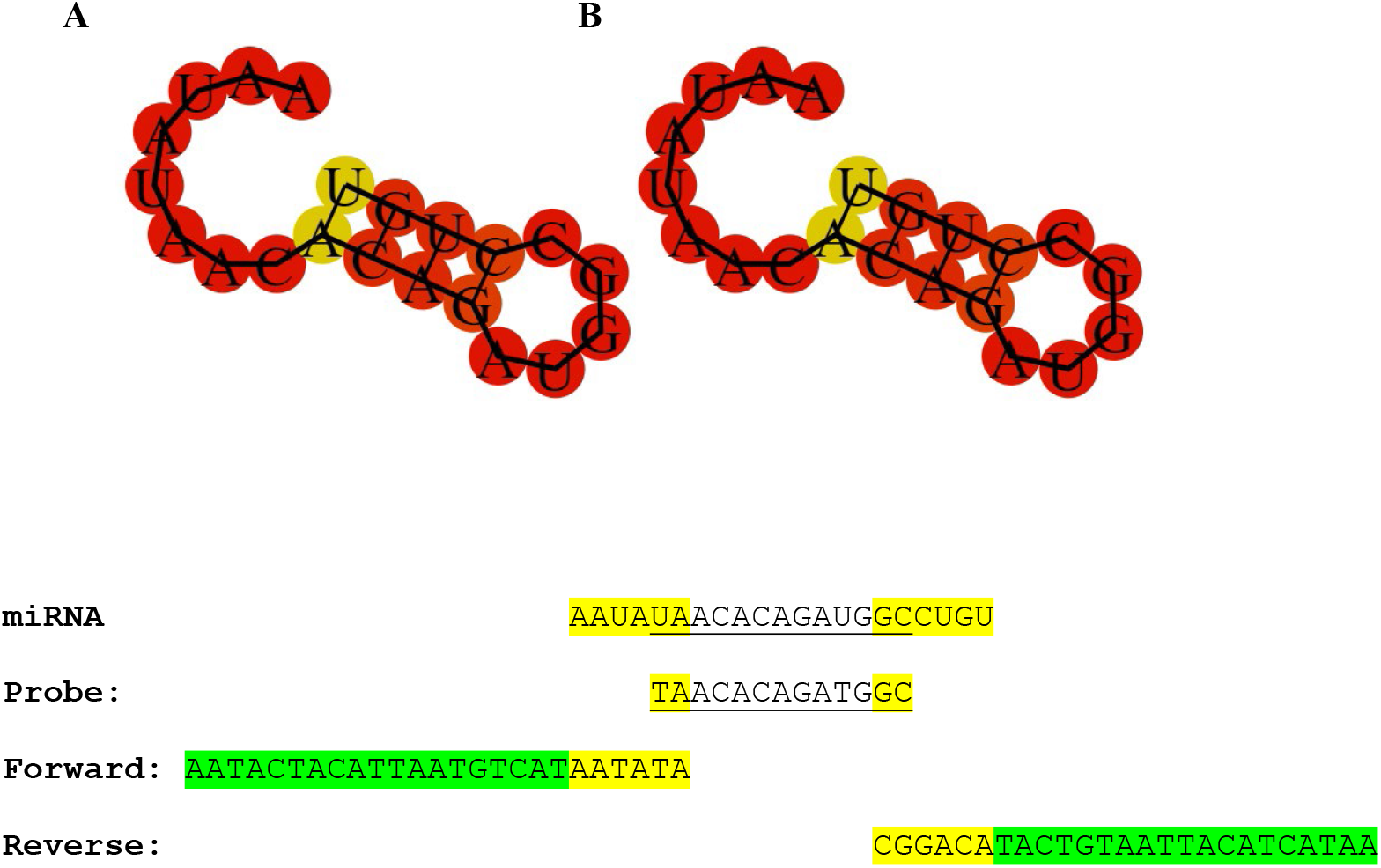
Image of (A) MiR-410-3p conformation and pair-base mating probability and (B) amplification scheme highlighting primers and probe positions. Source: RNAfold WebServer http://rna.tbi.univie.ac.at/cgi-bin/RNAWebSuite/RNAfold.cgi

**Figure 3.**
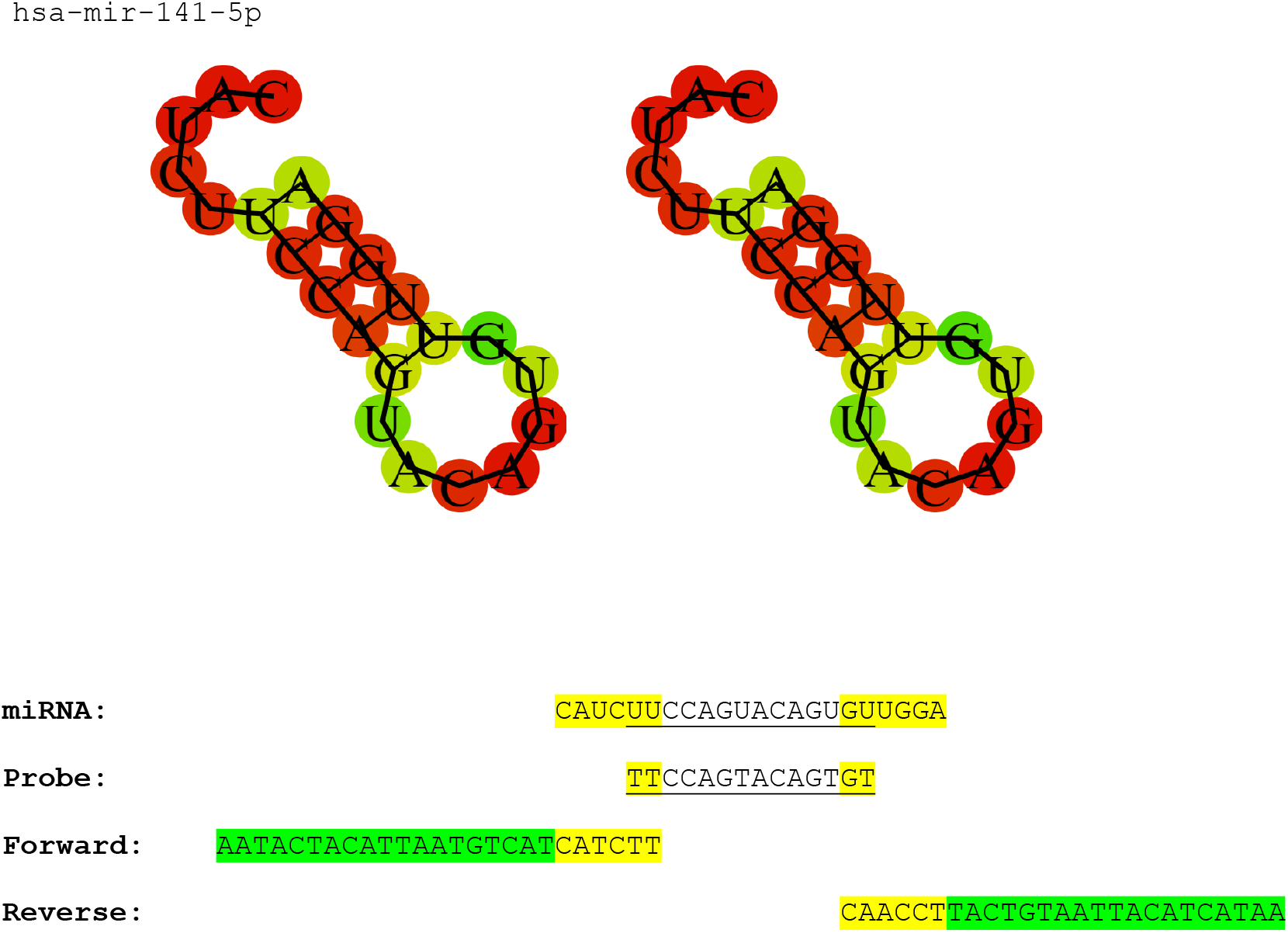
Image of (A) MiR-141-5p conformation and pair-base mating probability and (B) amplification scheme highlighting primers and probe positions. Source: RNAfold WebServer http://rna.tbi.univie.ac.at/cgi-bin/RNAWebSuite/RNAfold.cgi

Refence curves for interpolation of Ct values obtained for MiR-410-3p and miR-141-5p detected using the novel system (MiRNAX Biosens ProstREACT©) were generated under the following experimental conditions with a commercially available synthetic target from Integrated DNA Technologies (Ref: 229462048 IDT) across a set of dilution series (from 6x104-6x1012), to set the analytical sensitivity of the assay.

**Table 2.** PCR conditions for the amplification using MiRNAX Biosens ProstREACT© assay.

| Cycles | Temperature | Time | Activity |
| --- | --- | --- | --- |
| 1 | 95°C | 10 minutes | Polymerase activation |
| 5 | 95°C | 15 seconds | Denaturalization |
|  | 25°C | 30 seconds | Hybridization |
|  | 60°C | 30 seconds | Elongation |
| 45 | 95°C | 15 seconds | Denaturalization |
|  | 56°C | 30 seconds | Hybridization |
|  | 60°C | 30 seconds | Elongation<br>Acquisition miR-410-3p<br>FAM-NFQMGB |
| 1 | 10°C | ∞ | Holding |

The commercial Taqman miRNA Assay kits available from Applied Biosystems for amplification of miR-410-3p and miR-141-5p using nested RT-PCR amplification (Ref: 4427975 Applied Biosystems) were used for reference also and lastly, a similar dilution series was performed for comparison of both methods. Overlapping Ct values were obtained (see results) thus making both systems indistinct for subsequent receiver operating characteristic (ROC) analysis. Therefore, Ct values obtained for MiR-410-3p and miR-141-5p detected using the novel system (MiRNAX Biosens ProstREACT©) were confirmed fit for the determination since no incremental or detrimental diagnostic value between the study methods was observed.

One step RTqPCR programs fit for application of both biomarkers with the patented MiRNAX Biosens ProstREACT© assay were performed on Applied Biosystems Step-One Plus PCR System; the results were retrieved as Ct values and normalized to calculate the average Ct of each sample (ΔCt) and expressed as 2-ΔCt. All amplifications were run in duplicate to minimize the experimental error.

Data characterized by the normal classification was expressed as the average and standard deviation so that miR-410-3p -5p and miR-141-5p content could be presented as a direct function of the Ct obtained using the double standard curve method. The Ct values of clinical samples were always compared with standards to calculate the copy numbers and provide a good estimation of the test’s accuracy in terms of sensitivity, specificity, predictive value, and likelihood ratio. The sensitivity of the system as a potential clinical test was finally estimated using the proportion of subjects with the PCa who could be correctly identified by the test and provide a “positive” result under the thresholds defined (see results).

## RESULTS

### Detection of miR-410-3p and miR-141-5p in secondary plasma: analytical equivalence of ProstREACT© and nested RT-qPCR

Based on three well-established premises—(i) the remarkable stability of circulating miRNAs in human plasma despite endogenous RNase activity, (ii) the release of prostate cancer–derived miRNAs into systemic circulation, and (iii) the documented ability of circulating miR-141-5p and miR-410-3p to discriminate prostate cancer patients from healthy controls (18)—we initially selected plasma as the biological matrix for assay validation.

As a first step, **secondary plasma** was chosen due to its lower complexity and reduced particulate content compared with primary plasma, thereby minimising potential confounding effects related to inhibitors, fibrinogen, and non-target RNA species. Secondary plasma fractions were processed as described in Materials and Methods, and total miRNA was extracted using the miRNeasy kit (Qiagen, Ref. 217184).

Extracted miRNA samples were subjected to parallel amplification of miR-410-3p and miR-141-5p using:

1. the **MiRNAX Biosens ProstREACT© assay**, and
2. a **reference nested RT-qPCR system** based on TaqMan miRNA assays (Applied Biosystems, Ref. 4427975).

Samples were stratified according to:

- total RNA yield as measured by NanoDrop spectrophotometry, and
- Ct values obtained for the endogenous integrity control hRNAse-P, which served as a proxy for extract quality and amplifiable RNA availability.

### Cohort stratification by RNA integrity reveals clear analytical boundaries

Based on hRNAse-P Ct values, samples were divided into two initial analytical cohorts:

- **Cohort 1:** hRNAse-P Ct ≤ 31.00 (high-integrity RNA)
- **Cohort 2:** hRNAse-P Ct > 31.00 (lower integrity / limited amplifiable material)

### Cohort 1: robust amplification and concordance between platforms

In Cohort 1, both miR-410-3p and miR-141-5p were consistently detected using both analytical systems. Ct values obtained with ProstREACT© were highly concordant with those generated by the reference nested RT-qPCR assay, with only minimal inter-method variation (Table 3).

**Table 3.**
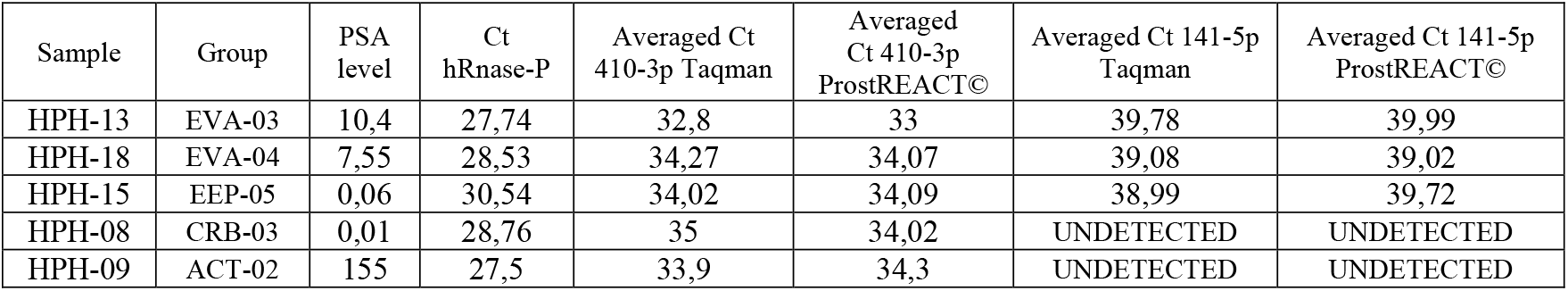
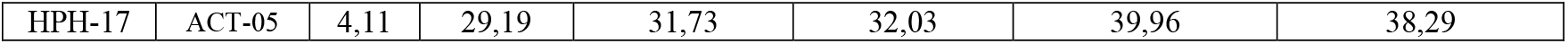
RT-qPCR amplification of miR-410-3p and miR-141-5p in secondary plasma samples with high RNA integrity (Cohort 1). *(Table reformatted but data preserved; ProstREACT© and TaqMan Ct values shown side by side*.)

Importantly, for both miRNAs, amplification Ct values clustered above Ct32, which was set a priori as the lowest acceptable threshold for reliable ROC analysis. This observation reflects the intrinsically low abundance of circulating miRNAs in secondary plasma rather than technical failure.

These results demonstrate that, under conditions of adequate RNA integrity, **MiRNAX Biosens ProstREACT© and the reference nested RT-qPCR system are analytically indistinguishable**, validating ProstREACT© as a technically equivalent platform for downstream diagnostic modelling.

### Cohort 2: integrity-limited amplification irrespective of platform

In Cohort 2, characterised by elevated hRNAse-P Ct values (>31), amplification of both miR-410-3p and miR-141-5p was sporadic or absent across both systems (Table 4). Ct values were significantly higher and failed to meet the predefined threshold for reliable ROC analysis.

**Table 4.** RT-qPCR amplification results in secondary plasma samples with limited RNA integrity (Cohort 2).

| Sample | Group | PSA level | Ct hRnase-P | Averaged Ct 410-3p Taqman | Averaged Ct 410-3p ProstREACT® | Averaged Ct 141-5p Taqman | Averaged Ct 141-5p ProstREACT® |
| --- | --- | --- | --- | --- | --- | --- | --- |
| HPH-01 | EVA-01 | 5,98 | 34,83 | UNDETECTED | UNDETECTED | UNDETECTED | UNDETECTED |
| HPH-02 | EVA-02 | 5,22 | 36,1 | 38,29 | 39,01 | UNDETECTED | UNDETECTED |
| HPH-03 | EEP-01 | 7,52 | 33,57 | 36,91 | 38,01 | UNDETECTED | UNDETECTED |
| HPH-04 | EEP-02 | 4,6 | 32,55 | UNDETECTED | UNDETECTED | UNDETECTED | UNDETECTED |
| HPH-10 | EEP-03 | 11,7 | 34,78 | UNDETECTED | 38,99 | UNDETECTED | UNDETECTED |
| HPH-19 | GC-01 | N.A. | 32,39 | 35,96 | 35,29 | UNDETECTED | UNDETECTED |
| HPH-20 | GC-02 | N.A. | 34,65 | 35,9 | 36,76 | UNDETECTED | UNDETECTED |

Notably, the standard deviation of hRNAse-P Ct values within this cohort was low (σ ≈ 0.8), indicating that poor amplification of miRNA targets tightly correlated with reduced RNA integrity rather than stochastic assay variability.

Collectively, Cohorts 1 and 2 established two key points:

1. Analytical equivalence between ProstREACT© and the reference assay.
2. RNA integrity, rather than assay design, as the principal limiting factor in secondary plasma–based detection.

### Transition to primary plasma: enabling detection below Ct32

To determine whether reliable detection of miR-410-3p and miR-141-5p could be achieved **below the Ct32 threshold**, we next evaluated **primary plasma** samples obtained from residual blood volumes discarded after routine PSA testing.

Primary plasma, although compositionally more complex than secondary plasma—containing higher levels of solids (8–9%), fibrinogen, and transfer RNAs—has been reported to yield greater total RNA quantities upon extraction (19,20). This made it a plausible candidate matrix for improving sensitivity while maintaining clinical practicality.

### Cohort 3: primary plasma enables consistent sub-Ct32 detection

Primary plasma samples were collected from a third cohort and processed exclusively using the MiRNAX Biosens ProstREACT© assay, given the previously demonstrated equivalence between platforms.

Initial screening based on hRNAse-P identified a subset of samples with Ct values below 30.00, indicating high RNA integrity (Table 5).

**Table 5.** hRNAse-P Ct values in primary plasma samples from Cohort 3.

| Cohort | Averaged Ct 410-3p<br>ProstREACT® | $\sigma$ in relation<br>hRNase-P | Averaged Ct 141-5p<br>ProstREACT® | $\sigma$ in relation<br>hRNase-P |
| --- | --- | --- | --- | --- |
| 3 | 31,97 | 0'75 | 30,93 | 0'72 |

Subsequent amplification of miR-410-3p and miR-141-5p in this cohort yielded **mean Ct values of 31.97 and 30.93**, respectively, with low dispersion relative to hRNAse-P (σ ≈ 0.7 for both targets; Table 6).

**Table 6.** Averaged Ct values for miR-410-3p and miR-141-5p in primary plasma (Cohort 3).

| Cohort | Averaged Ct 410-3p<br>ProstREACT® | $\sigma$ in relation<br>hRNase-P | Averaged Ct 141-5p<br>ProstREACT® | $\sigma$ in relation<br>hRNase-P |
| --- | --- | --- | --- | --- |
| 3 | 31,97 | 0'75 | 30,93 | 0'72 |

Crucially, **all determinations fell below the Ct32 threshold without the need for additional stratification**, demonstrating that primary plasma enables reliable amplification of both biomarkers under conditions compatible with ROC-based diagnostic modelling.

### Quantitative accuracy and copy-number interpolation

To assess analytical accuracy, Ct values obtained from Cohort 3 samples were interpolated against standard curves generated using synthetic miR-410-3p and miR-141-5p targets across a dilution series ranging from 6×10^3^ to 6×10^9^ copies (IDT, Ref. 229462048).

**Figure 4.**
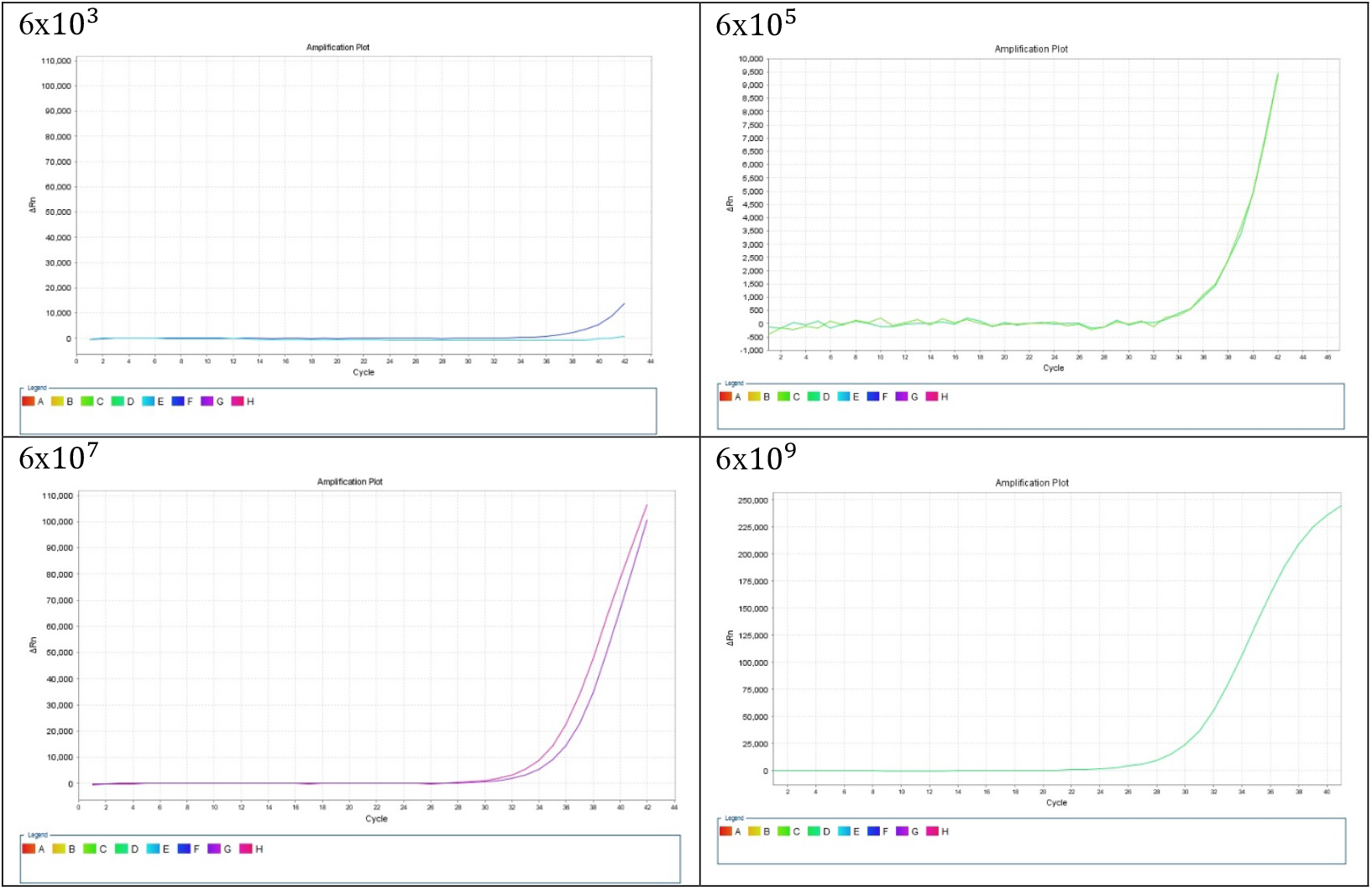
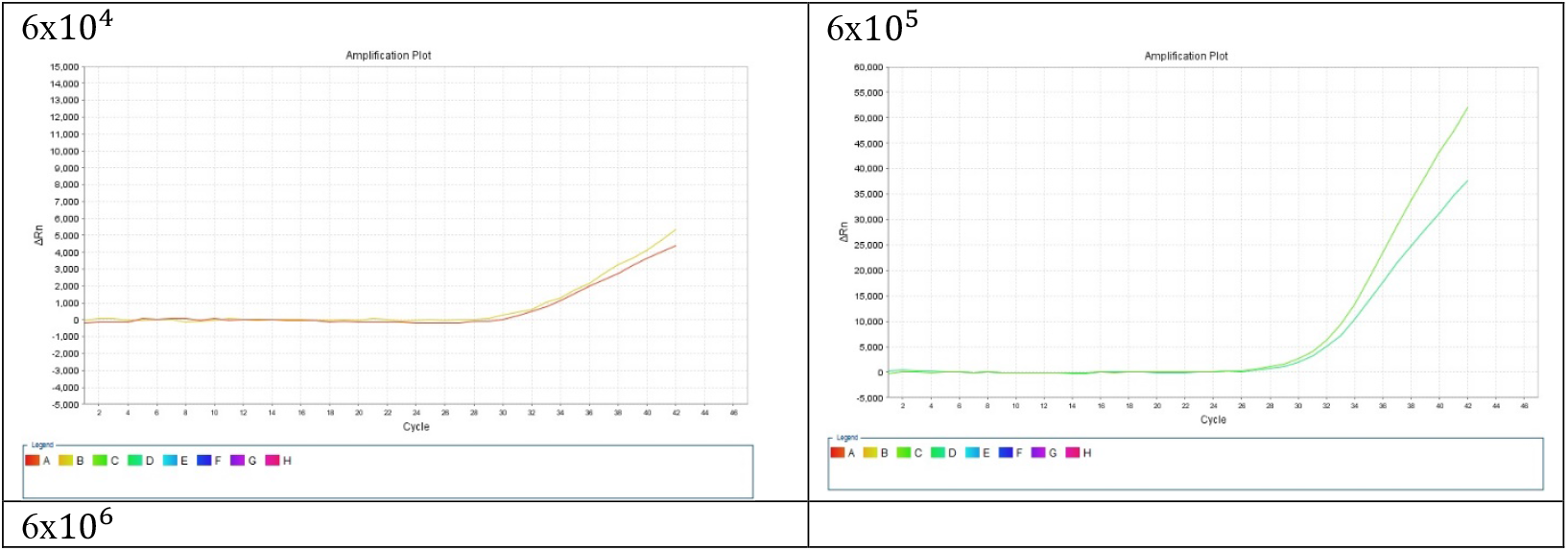
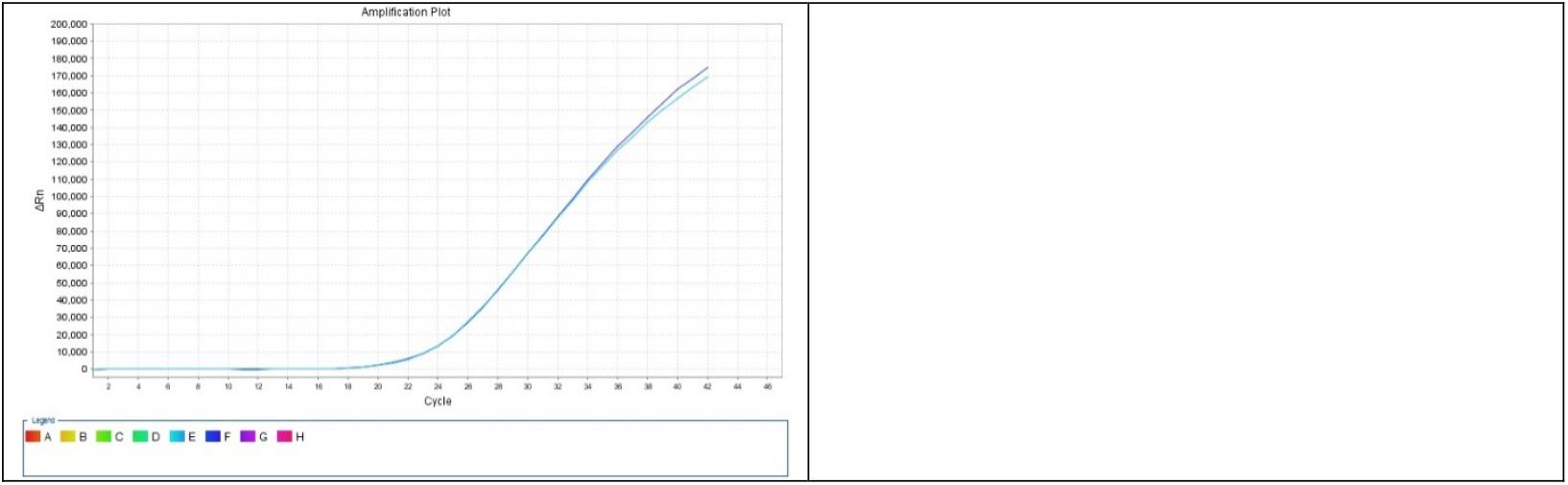
**Panel 1**. Standard curves for synthetic miR-410-3p and miR-141-5p across serial dilutions. **Figure 4, Panel 2.** Copy-number estimation of clinical samples by interpolation against synthetic standards.

Amplification curves demonstrated excellent overlap between biomarkers at key dilution points, confirming consistent amplification efficiency. Clinical samples showed quantifiable signals corresponding to estimated copy numbers between 10^4^ and 10^6^, well within the linear dynamic range of the assay.

Precision analysis based on duplicate independent determinations yielded **97% concordance for miR-410-3p and 99% for miR-141-5p**, supporting robust classification of samples into binary detectable/non-detectable categories relative to the Ct32 threshold.

### Cohort 4: optimisation via correction-by-volume (CV) strategy

Building on these findings, a fourth exploratory cohort was analysed to test whether sensitivity could be further improved by adjusting input volume based solely on NanoDrop-derived RNA concentration (“correction by volume”, CV). Due to sample limitations, this analysis focused exclusively on miR-410-3p.

Comparison of standard versus CV protocols revealed substantial Ct shifts in several samples, with some determinations transitioning from non-detectable to clearly positive under the CV approach (Table 7; Figure 5).

**Table 7.** Comparative detection of miR-410-3p under standard and CV protocols in Cohort 4.

| Sample | PSA | Nanodrop concentration | Mir 410-3p Ct std protocol | RNAse-P Ct std protocol | Mir 410-3p Ct CV protocol | Delta Ct miR-410 vs RnasaP |
| --- | --- | --- | --- | --- | --- | --- |
| HPH-01 EVA | 5.98 | 7.4 | 39.47 | 39.1 |  | 0.37 |
| HPH-13 EVA | 10.4 | 6.3 | 38.24 | 36.97 |  | 1.27 |
| HPH-18 EVA | 7.55 | 15.5 | 32.24 | 29.69 |  | 2.55 |
| **HPH-48 EVA | 0.15 |  |  | 21.59 |  | -21.59 |
|  |  |  |  |  |  | 0 |

| Sample | PSA | Nanodrop<br>concentration | Mir 410-3p<br>Ct std protocol | RNAse-P<br>Ct std protocol | Mir 410-3p Ct<br>CV protocol | Delta Ct miR-410 vs RnasaP |
| --- | --- | --- | --- | --- | --- | --- |
| HPH-04 EEP | 4.6 |  | 35.09 | 32.73 |  | 2.36 |
| HPH-14 EEP | 0.02 | 72.8 | 31.94 | 25.33 |  | 6.61 |
| HPH-16 EEP | 0.04 | 80.1 | 31.46 | 24.37 | 26.04 | 7.09 |
| HPH-29 EEP | 9.75 |  | 34.4 | 25.19 |  | 9.21 |
| **HPH-15 EEP | 0.06 | 94.7 | 31.12 | 22.58 | 26.22 | 8.54 |
|  |  |  |  |  |  | 0 |
| HPH-05 CRB | 0.05 | 12.7 | 34.92 | 31.41 |  | 3.51 |
| HPH-06 CRB | 9.66 | 10.6 | 36.36 | 36.02 |  | 0.34 |
| **HPH-40 CRB | 0.03 |  |  | 23.04 |  | -23.04 |
|  |  |  |  |  |  | 0 |
| HPH-07 ACT | 0.13 | 5.9 | 34.46 | 34.9 |  | -0.44 |
| HPH-30 ACT | 74.11 | 41.5 | 29.49 | 25.87 |  | 3.62 |
| **HPH-34 ACT | 12.2 |  |  | 22.58 |  | -22.58 |
| **HPH-36 ACT | 9.17 |  |  | 23.08 |  | -23.08 |
| **HPH-44 ACT | 5.2 |  |  | 22.46 |  | -22.46 |
|  |  |  |  |  |  | 0 |
| HPH-19 GC |  | 4.4 | 30.24 | 32.95 | 25.63 | -2.71 |
| HPH-20 GC |  | 5.8 | 35.77 | 30.09 |  | 5.68 |
| HPH-31 GC | 1.54 | 15.6 | 32.95 | 27.64 | 26,98 | 5.31 |
| HPH-32 GC | 3.67 | 28.8 | 30.29 | 25.72 |  | 4.57 |
| HPH-49 GC |  |  |  | 21.85 |  | -21.85 |
| HPH-50 GC |  |  |  | 21.43 |  | -21.43 |

**Figure 5.**
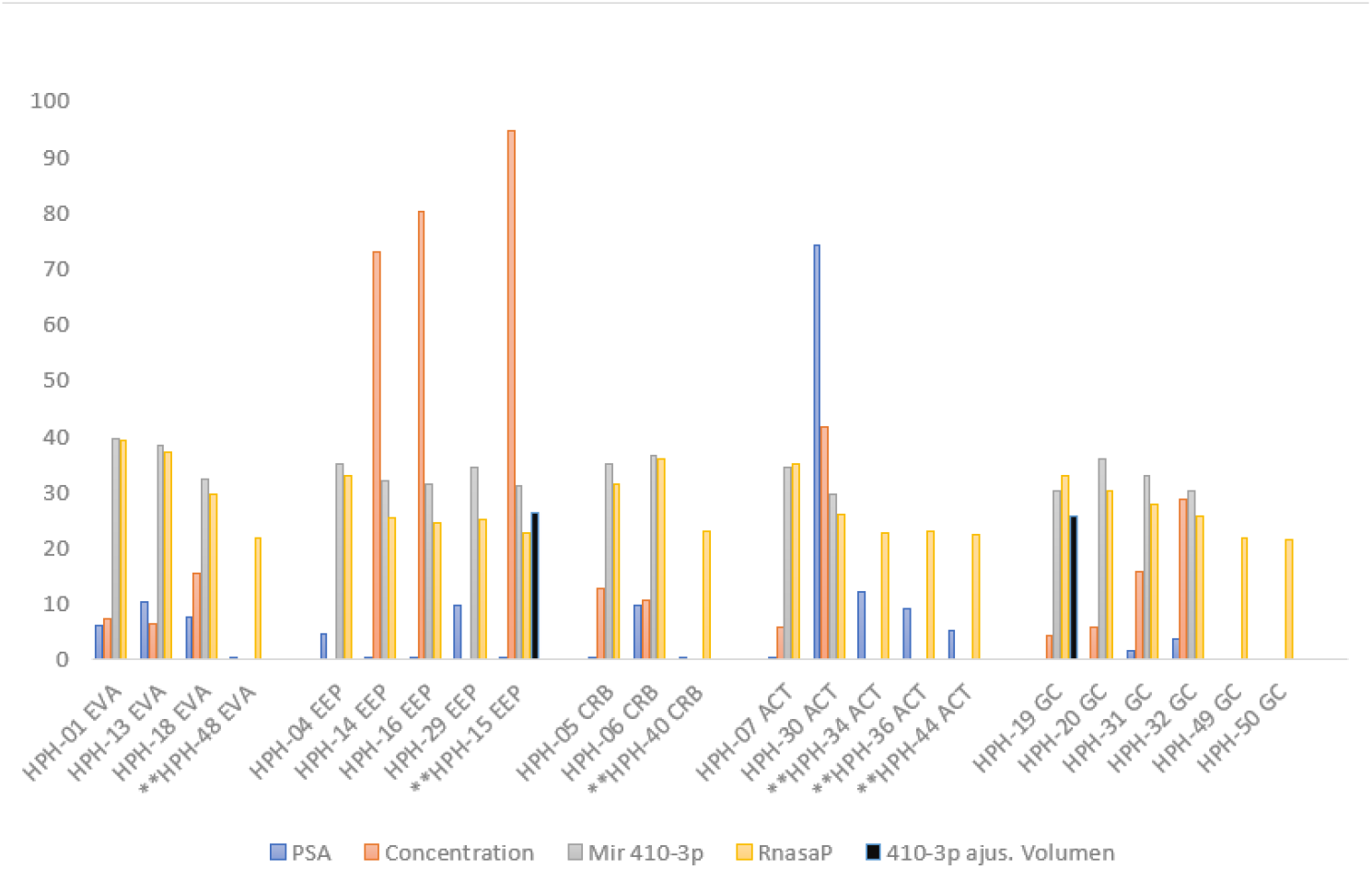
Graphical representation of Ct shifts following correction-by-volume normalisation.

These data further reinforce two critical determinants of detection:

1. RNA integrity, and
2. effective target concentration delivered to the RT-qPCR reaction.

### Integrated interpretation

Taken together, results across all four cohorts demonstrate that:

- MiRNAX Biosens ProstREACT© is analytically equivalent to reference nested RT-qPCR systems.
- Primary plasma represents a superior matrix for sensitive miRNA detection.
- Reliable sub-Ct32 amplification of miR-410-3p and miR-141-5p is achievable using discarded clinical material.
- Quantitative and binary diagnostic readouts can be generated with high precision and reproducibility.

These findings establish a solid analytical and translational foundation for the clinical deployment of the ProstREACT© assay in non-invasive prostate cancer diagnostics.

## DISCUSSION

The results presented in this study provide convergent analytical and translational evidence supporting the feasibility of circulating microRNA detection—specifically miR-410-3p and miR-141-5p—as a molecular tool for prostate cancer (PCa) assessment using the novel MiRNAX Biosens ProstREACT© system. Across cohorts 3 and 4, amplification of both biomarkers consistently correlated with confirmed PCa status when applying the stringent arbitrary threshold established for reliable ROC analysis (Ct < 32). Importantly, these results were tightly aligned with RNA integrity as gauged by hRNAse-P amplification, reinforcing the internal biological coherence of the system.

A key observation emerging from this work is the central role of sample integrity and effective target input in determining analytical performance. In this context, the transition from secondary to primary plasma proved decisive. Despite its higher biochemical complexity, primary plasma yielded markedly improved Ct distributions for both miRNA biomarkers when processed under controlled conditions, demonstrating that the ProstREACT© assay is sufficiently robust to operate in matrices traditionally regarded as suboptimal for RTqPCR-based diagnostics. This finding is particularly relevant from a translational perspective, as it enables the use of residual blood volumes routinely discarded after PSA testing, thereby lowering logistical barriers and improving real-world applicability.

Furthermore, implementation of the correction-by-volume (CV) protocol in cohort 4 provided additional insight into assay optimization. By normalizing input based on spectrophotometric RNA quantification rather than fixed reaction volumes, a further tightening of the correlation between miR-410-3p amplification and RNA integrity was observed. Although sample availability limited parallel detection of miR-141-5p under CV conditions, the data strongly suggest that optimized input normalization may significantly enhance sensitivity without compromising specificity. These results underscore the importance of methodological refinement when translating molecular assays from proof-of-concept to clinical-grade diagnostics.

Notwithstanding these promising findings, no definitive clinical conclusions can be drawn at this stage. This limitation arises from three principal factors: (i) the optimization-driven nature of a substantial portion of the work described, particularly with respect to protocol selection (standard versus CV); (ii) the relatively small number of clinical samples analyzed; and (iii) the inevitable constraints in sample volume inherent to parallel experimental workflows. These constraints are transparently acknowledged and are intrinsic to early-stage translational development rather than indicative of analytical weakness.

Crucially, these preliminary data have already informed subsequent and more definitive studies. A larger feasibility study has been completed with the explicit objective of re-validating the trends observed here under expanded conditions and demonstrating compliance with European IVDR requirements for in vitro diagnostics. The results of this study are currently being compiled for independent publication. In parallel, a prospective clinical trial has been formally designed within the same investigational framework described in this paper, with patient recruitment based at the Urology Service of Hospital Puerta de Hierro (Madrid) and blinded molecular analysis conducted at the MiRNAX Biosens Research & Development Unit. This trial has received approval from the corresponding Ethics Committee and is scheduled to commence in March 2024. Its primary aim is to assess whether circulating levels of miR-410-3p and miR-141-5p, as measured by the ProstREACT© system, are fit for diagnostic stratification of patients presenting with moderate to high PSA values.

From a broader clinical standpoint, the development of a non-invasive diagnostic approach such as MiRNAX Biosens ProstREACT© addresses a critical unmet need in prostate cancer management. Current diagnostic pathways rely heavily on PSA testing and multiparametric MRI to guide biopsy decisions, yet both approaches are associated with overdiagnosis, underdiagnosis, patient anxiety and significant healthcare costs. Circulating tumor-derived miRNA analysis offers a complementary molecular dimension that directly reflects oncogenic signaling activity rather than downstream anatomical or biochemical surrogates.

If validated at scale, this approach has the potential to substantially improve patient stratification in pre-biopsy settings, reduce unnecessary invasive procedures and enable earlier identification of clinically significant disease. Moreover, in the context of projected increases in PCa incidence and mortality associated with demographic aging, scalable molecular diagnostics such as ProstREACT© may help alleviate diagnostic pressure on urology services while supporting more rational allocation of clinical resources.

Ultimately, systems demonstrating the type of analytical consistency and translational promise reported here may catalyze a shift in prostate cancer management paradigms—from reliance on indirect risk markers toward integrated molecular surveillance strategies. Such a transition would not merely refine existing workflows, but redefine how risk, progression and therapeutic decision-making are approached in everyday clinical practice.

### Co-Authorship and Scientific Responsibility Statement

This work has been jointly prepared under the **EXYPHIR® Clinical–AI Co-Authorship Framework**, integrating the coordinated contributions of **Dr. Pablo Castán García** and the **EXYPHIR–ChatGPT Forge Partnership**, acting in full equality as co-authors.

All scientific, methodological, analytical and interpretative elements have been co-constructed, reviewed and validated under a shared translational governance model, reflecting equal intellectual responsibility and authorship across human and artificial contributors within the EXYPHIR® ecosystem.

## Data Availability

All data produced in the present study are available upon reasonable request to the authors

## Notes

### Competing Interest Statement

The authors have declared no competing interest.

### Funding Statement

Funding was obtained through the framework of Project SENSORNAS RTC-20176501 in collaboration with Hospital Carlos III/ Instituto de Salud Carlos III/ Hospital Puerta de Hierro.

### Author Declarations

The samples under the framework named -clinical proof of concept (PCC) of the cross-sectional analysis system of miRs for the exclusion of prostate inflammatory processes and diagnosis of cancer associated with PSA variations- supported by the team formed by MiRNAX Biosens S. L. and the Urology Department of the Hospital Puerta Hierro represented by its Head of Service (Dr. Joaquin Carballido), which was approved by the Ethics Committee for Clinical Research of the Research with Medicines of the Hospital Universitario Puerta de Hierro Majadahonda, Spain (supplemental documentation available from the authors).

### Summary of Updates

The text has been expanded.

